# Comprehensive analysis of *de novo* variants across 2,497 orofacial cleft trios reveals novel genetic drivers of disease

**DOI:** 10.64898/2026.05.21.26352934

**Authors:** Nehir E. Kurtas, Alba Sanchis-Juan, Eren Shin, Sarah W. Curtis, Kelsey R. Robinson, Arthur S. Lee, Azeez A. Alade, Xuefang Zhao, Jack Fu, Kimberly K. Diaz Perez, Lord J. J. Gowans, Mekonen A. Eshete, Wasiu L. Adeyemo, Carmen J. Buxó, Carmencita D. Padilla, Fernando A. Poletta, Angel Carreno Torres, George L. Wehby, Jacqueline T. Hecht, Lina M. Moreno Uribe, Nandita Mukhopadhyay, John R. Shaffer, Seth M. Weinberg, Jeffrey C. Murray, Terri H. Beaty, Azeez Butali, Michael Talkowski, Mary L. Marazita, Elizabeth J. Leslie-Clarkson, Harrison Brand

**Author notes:** **Corresponding author** Harrison Brand. These authors contributed equally to this work (Nehir E. Kurtas, Alba Sanchis-Juan).

## Abstract

**Background:** Orofacial clefts (OFCs) and other palate abnormalities (PAs) are among the most common birth defects worldwide and are characterized by the abnormal formation of the lip and/or palate. Genetic studies have traditionally classified OFC cases as either syndromic, involving OFCs alongside other congenital anomalies, or nonsyndromic, which represent the majority of cases and occur in isolation. Emerging genomic evidence indicates that genes traditionally associated with syndromic forms of OFC can also harbor variants contributing to isolated cases, challenging the notion of a strict dichotomy between these categories and supporting their integration for gene discovery.

**Methods:** In this study, we applied multiple analytic approaches to characterize the genetic architecture of OFC and PAs by integrating genomic data from 2,497 trios with probands diagnosed with an OFC (n=2,080) or PA (n=417). We compared these findings across OFC subtypes and syndromic status with those from 5,515 control trios to identify enriched biological pathways and mechanisms and to prioritize candidate genes using variant burden testing.

**Results:** We observed a significant enrichment of *de novo* protein-truncating and damaging missense variants in cases compared to controls (OR = 2.17, p = 1.21x10^-32^), with particularly strong signals in biologically relevant gene sets involving OFC-associated, constrained, Mendelian disorder, and mouse candidate genes. Variant burden testing identified 39 OFC risk genes at FDR ≤ 0.05, which we then integrated with 593 established OFC genes to interrogate the functional underpinnings of OFC via network analysis. This analysis revealed 309 high-order interactor genes not previously associated with OFC. Notably, this OFC network clustered into ten distinct biological pathways, with nucleosome-associated genes showing significant enrichment among cases in our cohort (OR = 14.8, p = 8.1x10^-4^). In a final integrative step, we combined evidence across all analyses to nominate 231 candidate genes, 32 of which contained at least two deleterious *de novo* variants in our cohort.

**Conclusions:** These findings underscore the value of integrating diverse OFC and PA subtypes, syndromic status, and variant classes to elucidate the genetic architecture of these disorders, highlighting both phenotypic expansion of known disease genes and the emergence of novel gene-phenotype associations.

## Background

Orofacial clefts (OFC) are characterized by the abnormal formation of the lip and/or palate during prenatal development and are among the most prevalent birth defects with a rate of ∼1 in 1,700 births [1]. Primarily, OFCs occur as three subtypes: cleft lip alone (CL), cleft lip with cleft palate (CLP), and cleft palate alone (CP) with each exhibiting distinct developmental trajectories, population prevalence, gender biases, and degrees of genetic heritability [2,3]. The genetic etiology of OFC subtypes appears to be somewhat distinct, with subtype-specific association signals that enrich different Gene Ontology (GO) terms [4]. OFCs are further classified as syndromic when they occur with additional congenital malformations (∼30% of cases), and nonsyndromic when they present in isolation (∼70% of cases) [5]. Although the biological mechanisms differentiating each subtype and syndromic forms are still poorly understood, recent studies have revealed a multifactorial genetic etiology, driven by a combination of common and rare variants. Currently, more than 45 loci have been discovered through genome wide association studies (GWAS) [6–10], and rare variants in more than 200 genes have been linked to Mendelian syndromes involving OFC [4,11,12]. Sequencing studies investigating genomic variants in OFC have highlighted a strong enrichment of *de novo* variants in both isolated and syndromic forms [4,13], and have demonstrated variants exhibiting variable expressivity and reduced penetrance, with rare pathogenic variants in genes associated with monogenic syndromes often identified in isolated forms of OFCs or other palate defects [3,14]. By example, variants in *IRF6* explain the majority of cases with Van der Woude syndrome (VWS) and Popliteal Pterygium syndrome (PPS) [15–17], yet *IRF6* variants are also significantly enriched in individuals with nonsyndromic OFCs [18].

Building on these prior findings, we have aggregated a large collection of exome (ES) and genome sequencing (GS) of 2,497 trio families with a proband affected with OFC and/or a palate abnormality (PA). Using this dataset, we characterize the distinct and shared genetic etiologies across OFC subtypes and syndromic status. To explore this, we employ complementary approaches, including gene-set enrichment analysis, rare-variant aggregation based gene discovery, and network analysis, to identify genes associated with OFCs, expand the phenotypic spectrum of known Mendelian disorder genes to include OFCs, and elucidate the biological mechanisms underlying OFC development. Overall, this study contributes to the growing body of literature demonstrating that rare variants play an important role in the genetic architecture of both syndromic and nonsyndromic OFC.

## Material and methods

### Sample Collection

Samples in our OFC and PA cohort were collected from 11 sources (Additional file 1: Table S1) and with each case including following inclusion criteria based on OFC or PA related Human Phenotype Ontology (HPO) terms (Additional File 2: Fig. S1). Samples with one or more HPO terms unrelated to OFC or PA were categorized as syndromic (Additional file 1: Table S2). We applied comprehensive quality control procedures to maintain data integrity across all samples in our study. For sex inference, we used a customized approach based on Somalier’s sex chromosome ploidy estimation method. Samples with inconsistent sex (n=7) were excluded from further analyses [19]. We applied the PC-Relate method, principal component analysis (PCA)-based method [20], across 17,766 known polymorphic sites to evaluate relatedness across the entire cohort to remove inconsistent family relationships, address sample swaps, and remove duplicate samples that can occur across multiple studies. Briefly, we computed pairwise kinship coefficients and identity-by-descent (IBD) sharing metrics (IBD0, IBD1, IBD2) between all sample pairs across cohorts. Expected relationships defined in the pedigree file were compared with these empirical relatedness estimates to identify ambiguous relatedness, sample contamination, or pedigree errors. Samples with poor relatedness quality control metrics were marked in the pedigree files for manual review and exclusion before downstream analyses. Control samples (n = 5,515) without reported OFC were collected from healthy siblings across two existing autism studies, with GS and ES *de novo* variants respectively curated from An et al. [21] and Fu et. al. [22]. As described below, these samples served as appropriate controls, as variant calling was conducted using comparable methods to those applied to the case samples.

### *De Novo* Variant Calling

Single nucleotide variants (SNVs) and insertions/deletions (indels) were detected with the Genome Analysis Toolkit (GATK) [23] with separate filtering procedures for de novo variants calling ES and GS. The ES filtering pipeline consisted of multiple steps adapted from Fu et al [22] (further details in Additional file 3: Supplementary Methods). Briefly, variants were retained only if they passed Variant Call Format (VCF) quality filters and multiple genotype-level criteria, including depth, genotype quality, Phred likelihoods, sex-specific ploidy checks, allele balance, call rate (≥0.8), and Hardy–Weinberg equilibrium (p ≥ 1×10⁻¹²), with remaining variants annotated using non-neuro Genome Aggregation Database (gnomAD) exome allele frequency priors. Additional filtering was applied by removing variants if they had parental allele balance >0.05, proband allele balance <0.25, depth ratio <0.1 between child and parents (proband read depth divided by the sum of parental read depth). We excluded low quality calls with genotype quality (GQ) <25 as well as those with a p-value greater than 0.05, an allelic depth (AD) of the alternate allele (AD_alt) threshold ≥10, variant quality score recalibration (VQSLOD) thresholds of ≥-20 for SNVs and ≥-2 for indels. We applied a final set of hard filters, keeping only heterozygous calls in the proband, and removing de novo variants that had a cohort allelic frequency (AF) >0.005 or gnomAD AF (genomes v2.1.1) >0.001 (if present). After all filtering, one variant was selected per person per gene, prioritizing variants with more severe consequences. SNV/indel variants were annotated for functional consequence using the Ensembl Variant Effect Predictor (VEP) v112 [24].

We identified de novo SNVs and indels from GS data using a modified version of the pipeline previously described by An. et. al. [21] and Werling et. al. [25] (further details in Additional file 3: Supplemental Methods). Briefly, variants were filtered by removing those with cohort allelic count (AC) ≥20 or located in low-complexity regions, retaining loci where parents were homozygous reference and probands heterozygous, applying stringent SNV- and indel-specific quality thresholds (e.g., VQSR PASS, QUAL, SOR, ReadPosRankSum, QD, MQ, GQ, AB, and DP), requiring mean GQ ≥50, and excluding outlier samples with excessive variant counts after sex and relatedness quality check (QC). De novo variants were detected with the following pipeline: 1) we apply the TrioDeNovo [26] package with the minDQ parameter set to two; 2) a final round of hard filters is applied, only retaining variants that passed all of these conditions: homopolymer length≤10, VQSLOD present, cohort-specific AC ≤2 or cohort AF ≤0.005, proband PL(0/1) =0, gnomAD AF (genomes v3.1.2) ≤0.001 (if present). After aggregating cohorts, we removed variants with AC >5. Given the higher number of false positives for de novo indels relative to SNVs, we trained a Positive-Unlabeled Random Forest (PURF) model to further filter de novo calls. Overall, these procedures provided high quality de novo callsets for both ES and GS. For the final call set, we performed an additional round of outlier filtering. SNV outliers were identified and removed using the 95th percentile (Q95) and an interquartile range (IQR) as follows: 1) in ES, samples exceeding the upper threshold (Q95 + 1.5×IQR) were excluded, with no lower bound applied; 2) in GS, both upper and lower thresholds were defined (upper: Q95 + 1.5×IQR; lower: Q95 − 3×IQR). For the control cohort, the callset was re-annotated on the Google Cloud Platform (vep-annotate-hail-ht) to match our case cohort and in GS filtered at an AF less than 0.001 in gnomAD r3.1.2 (GNOMADG_AF) to better match our case cohort, whereas the ES dataset was already filtered at this frequency [22]. A final QC step involved removing extreme outlier samples defined by an upper threshold of Q95 + 1.5×IQR, which resulted in 10 (8 in ES, 2 in GS) samples being removed from our cohorts.

### *De Novo* Structural Variant Detection

*De novo* structural variants (SVs) in our case cohort were identified for the aggregation-based gene discovery analysis. SVs were identified using the GATK-SV pipeline [27] and GATK-gCNV for GS and ES respectively [28]. Briefly, GATK-SV (https://github.com/broadinstitute/gatk-sv) integrates calls from multiple algorithms using paired-end and split-read evidence including Manta [29], Wham [30], and Melt [31], as well as read-depth–based methods cn.MOPS [32] and GATK-gCNV [28]. Subsequent steps include joint genotyping and variant filtering using machine learning–based approaches. Large variants located within recurrent genomic disorder (GD) regions were identified using bedtools [33], and evaluated separately and incorporated to the final call set after visual inspection using read-depth plots. For ES data GATK-gCNV [28] was applied to detect copy number variants (CNVs). The variant call set was annotated using svtk [25,27] to assess predicted functional impact and to compare allele frequencies with the gnomAD SV database (v2.1). Additional inheritance and frequency-based filters were applied to identify de novo (gnomAD SV AF ≤ 0.001, AC ≤ 5) candidate SVs. GD calls were kept exempt from the frequency filtering. All candidate SVs were manually curated and visually inspected using normalized read-depth plots [34] and Integrative Genomics Viewer [35].

### Generation of Enrichment Gene Lists

Gene-based analysis in this study focused on 18,128 autosomal genes (Additional file 1: Table S3) that have been previously shown to be well captured across various exome platforms to minimize platform-specific and technology-specific biases [22]. We further assessed enrichment across four gene-sets within the reference genes described below:

1. OFC genes were compiled from a comprehensive list of 593 autosomal genes linked to OFC taken from three partially overlapping sources: (A) 383 established OFC genes curated by Diaz Perez et al. [11], (B) 349 genes from the HPO catalog filtered for relevance to OFC phenotypes [36], and (C) 117 genes derived from GWAS studies of cleft-related traits based on GWAS studies from the NHGRI-EBI GWAS catalog (v1.0 downloaded from https://www.ebi.ac.uk/gwas/; searched terms “cleft” or “palate” and then manually reviewed to ensure study was OFC related) [37]. Among the GWAS variants located at an intergenic site, we select the nearest gene.
2. We collected the human homologs of mouse genes associated with OFC from the Mouse genome informatics (MGI) database [38], and applied filtering to include relevant Mouse Phenotype terms, and retained 349 genes overlapping our above autosomal coding genes (further details in Additional file 3: Supplemental Methods).
3. Online Mendelian Inheritance in Man (OMIM) genes [12] were mapped to our global gene list and autosomal dominant (AD) annotations were assigned using the OMIM gene-phenotype mapping file; overall, 4,486 genes were mapped to OMIM, of which 1,928 had an OMIM AD annotation.
4. Genes under evolutionary constraint were considered haploinsufficient when they had a gnomAD v2 Loss-of-function Observed/Expected Upper bound Fraction (LOEUF) score ≤ 0.35) [39], totaling 2,746 constrained genes in our global gene list.

### Enrichment Analysis

*De novo* SNVs and indels from cases and controls were analyzed to assess mutational burden across reference gene sets, with annotated gene subsets as described above. Variants were classified into functional categories. Missense variants were further stratified into functional categories based on missense deleteriousness metric (MPC) [40]. Specifically, variants annotated as missense were classified as misB (highly disruptive) if MPC ≥ 2, misA (moderately disruptive) if 1 ≤ MPC < 2 and MisOther (low impact missense) if MPC < 1. Protein-truncating variants (PTVs) included variants with “frameshift_variant”, “splice_donor_variant”, “splice_acceptor_variant”, and “stop_gained” functional consequences. Gene enrichment analysis included PTV, misB, misA, MisOther, and synonymous variants. Carrier counts were computed per individual, ensuring that each sample contributed at most one variant per gene, and aggregated across gene sets and variant classes. Enrichment analyses were performed using two-sided Fisher’s exact tests by comparing the number of variant carriers and non-carriers between cases and controls. P-values were calculated for each variant class and gene set, and multiple testing correction was applied using the Bonferroni method.

### Identification of OFC Risk Genes

We performed gene-level association analysis with the transmission and *de novo* association (TADA) model. TADA is a Bayesian analytical framework for gene-level association studies [22,41,42]. Briefly, evidence of association is represented through a Bayes Factor (BF) that is calculated for each gene across different variants considering mutation rates and prior relative risk. The combined BFs generate a false discovery rate (FDR) based q-value with a lower q-value representing a stronger significance. We calibrated the model with existing mutation rates and relative risks from a previously published study [22]. We applied the model on five variant classes (1) PTV, (2) misB, (3) misA, (4) predicted loss-of function SV (SV LoF), and (5) predicted copy gain duplications. Significant genes were determined with an FDR<0.001, which we have shown to be equivalent to a Bonferroni correction [22].

### Network Analysis

We extracted protein-protein interactions using the STRING protein interaction database (version 12) [43] across 610 seed genes, comprising the previously defined OFC-associated gene list from above (n = 591; note MAPK10 and OFCC1 were excluded due to lack of matching annotation in the STRING database) and additional OFC risk genes derived from our TADA analysis at FDR ≤ 0.05 (n = 19). We rigorously classified interactions by combining probabilities of four selected evidence types (database, experiments, experiments transferred and database transferred), which were adjusted based on the chance of randomly observing an interaction (probability = 0.041), as recommended on the STRING website. We retained only interactions with a combined score of ≥0.9. We hypothesized more connected first-order interactor genes would have a stronger association with mutations in our cohort and we assessed this by binning OFC interactors based on their number of interactions with seed genes and tested for enrichment of de novo deleterious mutations (misB+PTV). Association between mutation status and bin membership was assessed using a Fisher’s exact test applied to counts of genes harboring PTVs and misB mutations in cases versus those without these mutations. Odds ratios, 95% confidence intervals, and two-sided p-values were computed, with Bonferroni correction applied for multiple testing corresponding to 6 interaction bins based on the number of edges per gene (edge 1, edge 2, edge 3, edge 4, edge 5, and edge >5) and 2 mutation categories (deleterious vs. synonymous). To extract the complete network for these 309 high order interactors and 610 seed genes, we used the STRING web service (https://string-db.org/, v12.0). The analysis is restricted to full STRING networks (network type), using experiments and databases as sources (active interaction sources) with an interaction score of more than 0.9 (high confidence). The resulting network was visualized using the Cytoscape app (v3.10.4) [44].

We sought to identify protein clusters with shared biological functions within our final network by applying a Markov Cluster Algorithm (MCL) [45] in STRING, using the default inflation parameter of 3. The algorithm uses the distance matrix produced from STRING global scores so that proteins with higher interaction scores are more likely to be grouped into the same cluster. Pathway enrichment analyses were conducted separately for 10 MCL derived clusters using the *clusterProfiler* package [46], with the background defined as 18,128 genes with successfully mapped Entrez IDs [47]. For each GO [48,49], enrichment was computed using a hypergeometric framework, and results were re-evaluated using one-sided (alternative=“greater”) Fisher’s exact tests. Finally, we assessed *de novo* enrichment in our cohort for each of the corresponding clusters encompassing 10 or more genes through a gene set enrichment as described above.

## Results

### Composition of the OFC and PA Cohort

We uniformly processed and harmonized sequencing data from 2,080 OFC case-parent trios, supplemented with 417 trios with PA (e.g., high or narrow palate) but no OFC, under the hypothesis these cases serve as a milder manifestation of CP and could therefore provide additional power to identify shared genetic risk factors. We leveraged both GS (n = 1,596 trios) and ES (n = 901 trios) data derived from 11 sources (Additional File 1: Table S1) with detailed phenotypic information provided in human phenotype ontology (HPO) format. The cohort encompasses the following phenotypic subtypes: CP (GS: 399; ES: 337), CL (GS: 190; ES: 60), CLP (GS: 796; ES: 82), an unspecified OFC type (uOFC) (GS: 192; ES: 24) and PA (GS: 19; ES: 398) (Additional file 1: Table S2). Mirroring known clinical demographics, the majority of probands had an isolated OFC absent of other anomalies (n = 1,586; 63.5%) at the time of recruitment [50,51], with the remainder classified as syndromic, indicating the presence of one or more additional phenotypic features beyond their cleft or palate defect. Among the syndromic cases, the cohort comprised a total of 2,219 distinct comorbidities with a median of 10 HPO terms per case and the most common terms involved the nervous and/or musculoskeletal systems (Additional File 2: Fig. S2). Among the entire cohort there was significantly higher number of male probands (n = 1,412) compared to females (n = 1,085) (binomial test, p = 6.45×10^-11^) and this was more pronounced between nonsyndromic cases (male:female ratio = 1.40) vs syndromic cases (male:female ratio = 1.14) (Fisher’s exact test, p = 0.02). These findings are consistent with epidemiologic studies showing more frequent occurrence of OFC in males vs females [52–54]. OFC substypes exhibited characteristic sex biases with CP observed more frequently in females (male:female ratio = 0.9), whereas CL and CL/P cases (male:female ratio = 1.8) occurred more frequently in males [55,56]. To supplement this cohort, we assembled a control cohort composed of unaffected siblings of individuals with autism spectrum disorder (ASD) from two published studies: 3,615 siblings with ES [22] and 1,900 siblings with GS (An et al., 2018). Importantly, these cohorts were processed using similar variant-calling pipelines and displayed comparable *de novo* rates for synonymous variants (binomial test, p = 0.62), which should be independent of disease architecture given their largely neutral effect on protein function (Additional File 2: Fig. S3).

Across this cohort, we collected 6,778 *de novo* non-synonymous PTV and missense SNVs/indels in patients and controls. Consistent with well-established patterns [4,13], these variants were enriched in probands, with 62.4% harboring at least one *de novo* mutation compared to 53.2% of controls (Chi-square test, p = 1.86x10^-32^). We further broke down our missense mutations based on the *in silico* Missense badness, PolyPhen-2, and Constraint (MPC) metric [40], partitioning these variants into three categories: misB (MPC>=2; highly disruptive), misA (2 > MPC >=1; moderately disruptive), and misOther (MPC < 1; low impact) [22]. Beyond short variants, we detected 435 *de novo* SVs in our probands across GS data using GATK-SV [27,57] and ES with GATK-gCNV [22,28]. The rate of *de novo* SVs observed in probands was approximately 0.18 (1 per 5.5 individuals) from GS and 0.08 coding CNVs from ES. Further grouping these SVs by subtypes, we observed 198 deletions (DEL), 161 duplications (DUP), 56 insertions (INS), 15 complex SVs (CPX), 3 reciprocal translocations (CTX) and 2 inversions (INV). Notably, due to insufficient control data with *de novo* SVs, we excluded SVs from the enrichment analyses below, but they were still included in the gene discovery component, which does not rely on controls.

### Enrichment Analysis

We initially examined the *de novo* mutational burden stratified by mutation and phenotype subtypes (Fig. 1A; Additional File 1: Table S4). We observed a striking enrichment of *de novo* deleterious mutations (PTVs, misB) across the aggregate OFC and PA cases (n=2,497) when compared against controls (Fisher’s exact test, OR=2.17, p=1.21×10^-32^), but no difference in cases compared to controls for synonymous variants (p = 0.8) or the less disruptive missense annotations (misA [p = 0.24], MisOther [p = 0.086]) (Fig. 1A). The enrichment of misB variants (Fisher’s exact test, OR=2.6, p=2.48×10^-18^) was higher than the enrichment detected for PTV (Fisher’s exact test, OR=1.8, p=3.71×10^-15^), though the misB score incorporates missense constraint information. Similarly, restricting analyses to the group of PTV mutations that undergo the strongest negative selective pressure using the loss-of-function observed/expected upper bound fraction metric [39] (LOEUF, score <0.35) revealed an even stronger enrichment of PTVs in cases (Fisher’s exact test, OR=4.8, p=1.56×10^-32^). Further subgrouping found that *de novo* deleterious variant enrichment was highest in syndromic OFC cases, with misB variants consistently showing the highest enrichment (Fisher’s exact test, OR=4.8, p=1.08×10^-19^) (Additional file 1: Table S2).

**Fig. 1.**
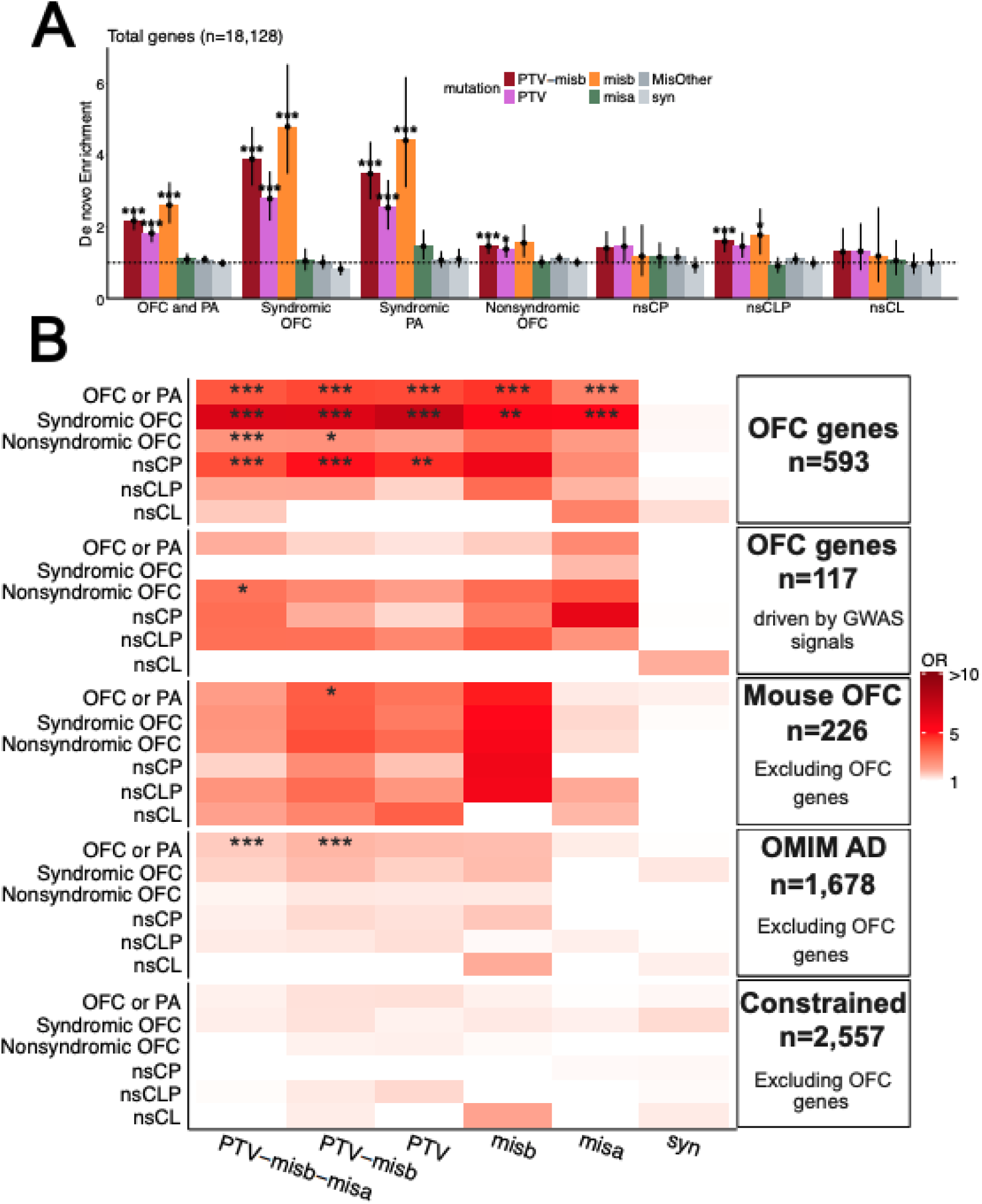
*De novo* variant enrichment across functional and clinically OFC relevant gene sets. **(A)** *De novo* enrichment analysis of autosomal coding genes (n = 18,128) stratified by phenotype and mutation type compared against 5,515 control trios. Phenotypic categories include OFC or PA, syndromic OFC, syndromic PA, nonsyndromic OFC, and nonsyndromic subtypes such as CP, CL, and CLP. Mutation categories include PTV, misB (MPC>=2; highly disruptive), misA (2 > MPC >=1; moderately disruptive), MisOther (MPC < 1; low impact), syn. Bars show an odd ratio for each test with a 95% confidence interval (CI) indicated by error bars. Null odds ratio of 1 is indicated with a horizontal dashed line. **(B)** Heatmap showing enrichment across relevant gene sets grouped by phenotypic subgroups (rows) and mutation classes (columns). Gene sets include: OFC genes (n = 593), OFC genes driven by GWAS loci (n = 117), mouse MGI OFC genes excluding known OFC genes (n = 226), OMIM AD genes excluding OFC genes (n = 1,678), and constrained genes excluding OFC (n = 2,557) (Additional file 1: Table S3). Color intensity is based on the magnitude of enrichment. Asterisks highlight statistical significance (*p < 0.05, **p < 0.01, ***p < 0.001) of p-values after Bonferroni correction.

We next examined whether the observed enrichment was preferentially concentrated within biologically or clinically relevant gene sets (Additional file 1: Table S3). Given the presence of a global *de novo* enrichment signal across all genes, we evaluated each gene set enrichment relative to this baseline expectation to determine whether excess burden was preferentially concentrated within these specific groups (Fig. 1B; Additional File 1: Table S5; unadjusted results can be found in Additional File 1: Table S5 as well). As expected, genes previously associated with OFC (n = 593) showed significant enrichment for *de novo* PTV or misB mutations even above the background enrichment (Fisher’s exact test, OR=4.2, p=1.08×10^-14^). This held true when subsetting to those with support from common variants derived from genome wide association studies (GWAS) (n = 117), particularly among the nonsyndromic cases, which comprise the majority of GWAS cohorts, though only when combining across PTV-misb-misa (Fisher’s exact test, OR=3.24,p=7.5×10^-4^) did the signal surpass the threshold for multiple testing correction (Fig. 1B; Additional File 1: Table S5). Overall, this highlights a convergence of rare and common variant signals in OFC as reported in smaller cohorts [4,58,59]. Conversely, genes previously implicated with rare variants in OFC (n=490) showed the highest enrichment of deleterious variants (Fisher’s exact test, OR=6.1, p=1.28×10^-18^) in the overall cohort.

We next investigated enrichment beyond established OFC genes and found a significant excess of *de novo* deleterious mutations (Fisher’s exact test, OR=1.8, p=1.7x10^-5^) in Mendelian disorder genes classified as AD in OMIM [12] but not observed in our OFC gene list (n = 1,678) (Fig. 1B, Additional File 1: Table S5). These findings suggest strong potential for phenotype expansion among many of these disorders as discussed below. Another source of biologically plausible novel OFC genes arises from mouse studies that have not yet been characterized in humans. We investigated such genes by parsing the MGI database and mapping the mouse data to their human orthologs. After removing established OFC genes, we observed 226 genes with a strong enrichment *de novo* enrichment (Fisher’s exact test, OR=3.75, p=6.9x10^-4^) (Fig. 1B, Additional File 1: Table S5). Finally, we explored enrichment within dosage-sensitive genes. We focused on 2,746 genes under evolutionary constraint for LoF mutations via the gnomAD v2 *LOEUF* model (*LOEUF* score ≤ 0.35) [39] and observed a *de novo* deleterious variant enrichment (Fisher’s exact test, OR=1.9, p=1.8x10^-9^) (Additional File 1: Table S5), which remained nominally significant after excluding established OFC genes (n=2,557) (Fisher’s exact test, OR=1.3, p=0.02).

### Gene Association

Moving beyond these enrichment analyses, we next sought to identify novel candidate genes through integrated association analyses [22,41]. TADA is a Bayesian framework that robustly integrates genetic evidence across variant classes (SNVs/indels [PTV, misA, misB] and SVs), study designs, inheritance modes, and sequencing modalities (i.e., genome and exome sequencing). Briefly, TADA evaluates gene-phenotype associations by comparing the likelihood of observed variant counts under a null model of no association with an alternative model that incorporates prior information from gene constraint metrics, such as LOEUF scores (see methods for further details). These analyses detected 39 genes at an FDR<0.05, 16 of which were significant at FDR<0.001, which approximates a genome-wide Bonferroni correction [22] (Fig. 2A; Additional File 1: Table S6). The majority of associations at FDR ≤ 0.05 were driven by PTV mutations (57%) compared to 36% from misB (Fig. 2B). Importantly the inclusion of SVs, which are often excluded in association studies due to analysis challenges, provided enough additional evidence to shift two genes (ZFHX3 and GRHL2) toward significance (FDR ≤ 0.05). We next investigated the phenotype of the cases contributing mutations to the association signal per gene (Fig. 2A), and detected a higher contribution of syndromic cases (69.3%) than nonsyndromic cases (Fig. 2C), but 50% of the genes had at least one nonsyndromic case contributing, consistent with variable expressivity of these genes.

**Fig. 2.**
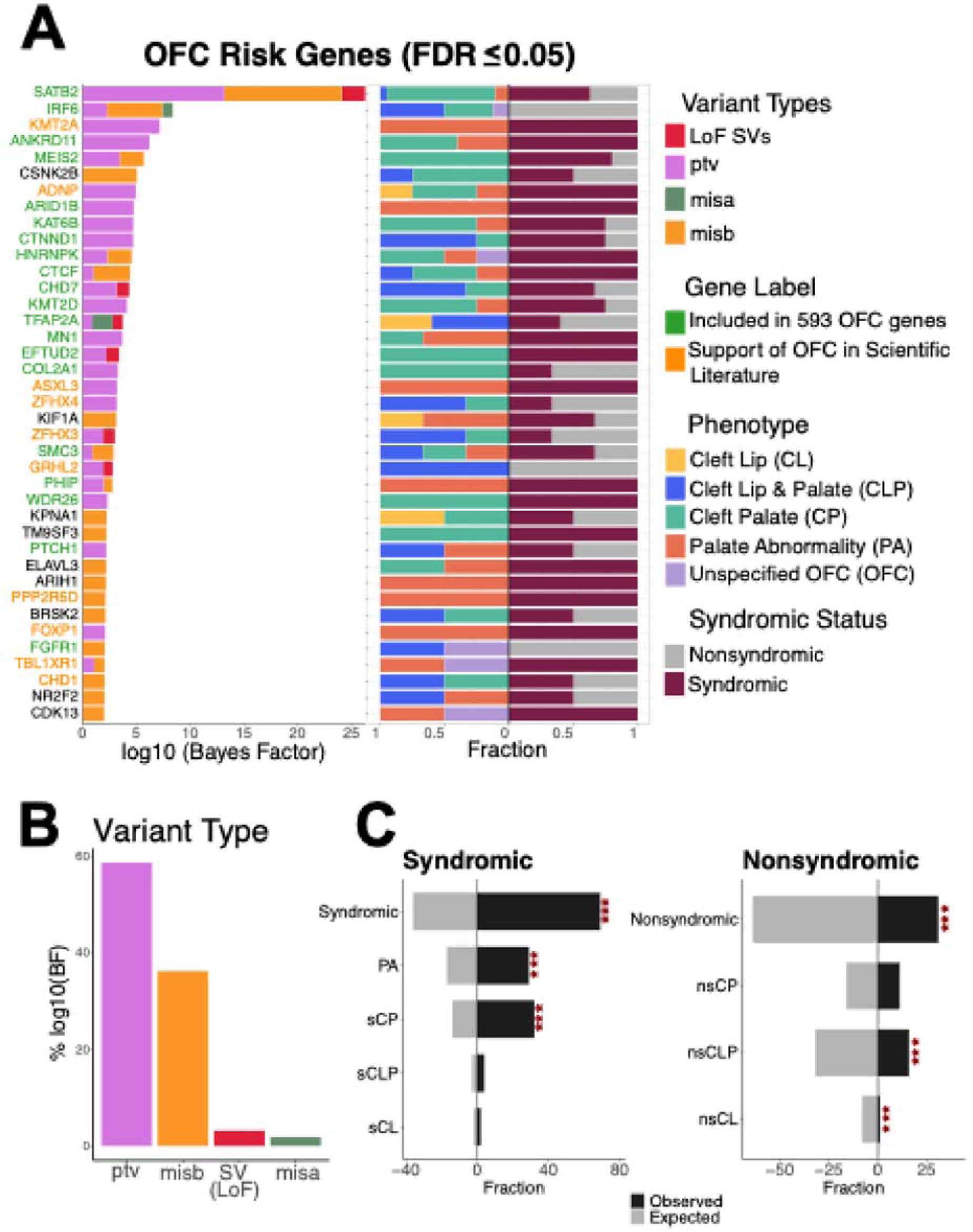
Gene association analysis. **(A)** Gene-level association results for TADA analysis, with significant genes at an FDR ≤ 0.05. The left panel shows the strength of association for each gene measured with log10 Bayes factors. Each bar is coloured based on the contribution of different variant classes, including structural variants and SNV/indels (LoF SVs, PTVs, misB and misA). Gene labels are annotated to demonstrate literature evidence of association: genes previously reported in OFC cohorts (in 593 gene list, green) and genes supported by the scientific literature (orange). In the right panel, each bar represents the fraction of samples contributing to the association signal for each gene based on either syndromic status or OFC/PA subtype. **(B)** Contribution of variant classes to the overall association signal across all significant genes at FDR ≤ 0.05. **(C)** Comparison of observed versus expected contributions of OFC/PA subtypes to the overall association signal. Asterisks highlight statistical significance of p-values from a binomial test (*p < 0.05, **p < 0.01, ***p < 0.001).

Further stratifying the 16 significant genes at an FDR<0.001, revealed 13 as established OFC genes with *SATB2* [60] showing the strongest association followed by *IRF6* [61]. Among the three remaining genes, each was supported by evidence in the literature implicating association with OFC. *KMT2A* is associated with Wiedemann-Steiner syndrome (MIM605130) which is characterized by developmental delay, intellectual disability, and characteristic facial appearance [62]. Case studies reported CP in a subset of patients with *KMT2A* mutations [63,64], and it was recently highlighted as a candidate gene for nonsyndromic OFCs based on proximity near a GWAS signal [65]. *ADNP* is associated with Helsmoortel-van der Aa syndrome (MIM615873), which is a neurodevelopmental disorder characterized by developmental delay, hypotonia and characteristic facial features with CP and CL/P findings reported in a subset of individuals [66,67]. Finally, we observed an enrichment of mutations in *CSNK2B,* which is associated with Poirier-Bienvenu neurodevelopmental syndrome (POBINDS, MIM618732) and characterized by developmental delay and early-onset epilepsy. While facial dysmorphisms are reported frequently for POBINDS cases, specific facial features are not as well established, though two cases have been reported as having CP [68]. Unlike *ADNP* and *KMT2A*, which were observed exclusively in syndromic cases, approximately half of the mutations identified in *CSNK2B* occurred in individuals classified as nonsyndromic, suggesting either an expansion of the phenotypic spectrum associated with this gene or a later disease onset with more subtle structural features that may have been missed during the initial syndromic assessment. Either way, this analysis adds to the growing body of genetic evidence supporting a role for this gene in OFC.

### Network Analysis

Recognizing that our sample size is still limited in gene discovery power, we sought alternative methods to prioritize potential OFC candidate genes. Building off orthogonal biological insights from protein-protein interaction (PPI) networks we investigated additional genes and pathways that may be associated with OFC. We constructed a STRING protein network using a seed list consisting of our 593 established OFC genes (Additional file 1: Table S3) and 19 additional significant genes from our TADA analysis described above (FDR<0.05). We hypothesized that greater connectivity among first-order interactor genes, defined as those not included in the seed list but connected to at least one seed gene, would be associated with increased burden of de novo mutations, and accordingly evaluated this enrichment (n=2,639; interaction score ≥0.9). Examination of each first-order interactor stratified by the number of connections with the 612 OFC seed genes showed those with at least five connections (n = 309) represent an inflection point for significant enrichment (Fisher’s exact test, OR=4, p=2.9x10-3, Additional file 2: Fig. S4). Accordingly, we constructed a high-confidence interconnected network comprising the 309 high-order interactors and 224 OFC seed genes they directly connect with (Fig. 3A).

**Fig. 3.**
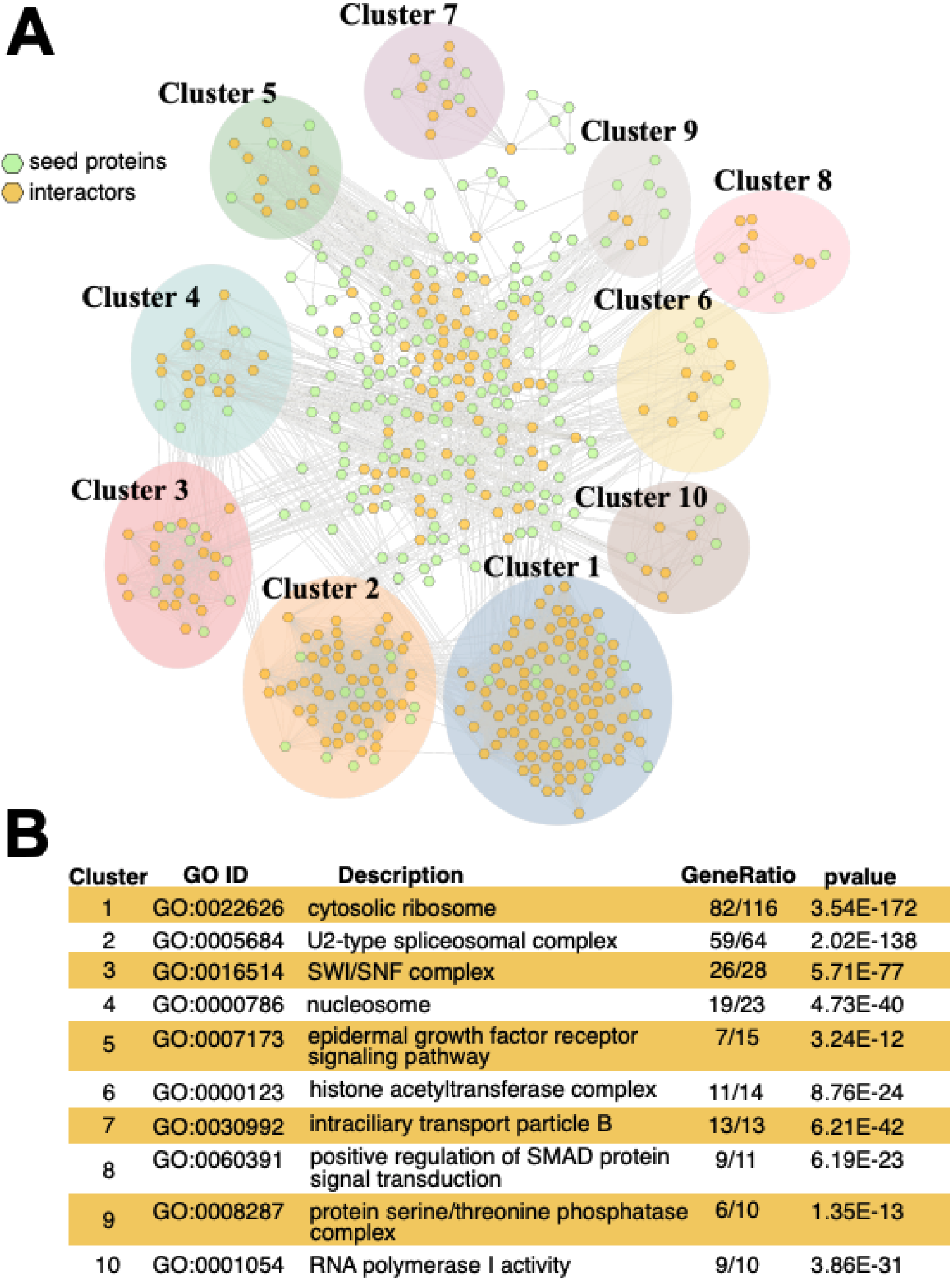
OFC protein interaction network. **(A)** Network representation of seed proteins (green), derived from OFC-associated gene lists, and their interactors (yellow), visualized using Cytoscape. Nodes were grouped into clusters using the Markov Cluster (MCL) algorithm. Each shaded region represents a distinct cluster, with the top 10 clusters (ranked by number of genes) highlighted. **(B)** GO enrichment analysis for each cluster. The most significantly enriched GO term per cluster, after Bonferroni correction, is shown along with the corresponding GO ID, description, gene ratio (number of genes in the cluster annotated with the term relative to the total number of genes in the cluster), and nominal p-value.

In order to explore OFC-related biological functions or pathways associated with our high-confidence network, we applied a MCL using STRING [43,45] and identified 81 total clusters. Among these, ten contained ≥10 genes, with the largest cluster consisting of 118 genes (Additional File 1: Table S7). To better understand potential shared functional mechanisms underlying these groups, we performed a pathway enrichment analysis for each of the 10 clusters using GO [48,49] (Additional File 1: Table S8. Pathways were found to be related to gene expression and RNA processing, including RNA polymerase I and spliceosomal complex, as well as chromatin organization pathways, such as nucleosome, histone acetyltransferase, and SWI/SNF complexes (Fig. 3B). We next assessed whether genes within specific clusters showed enrichment for *de novo* variants. We detected at least a nominal enrichment of *de novo* PTV or misB enrichment in clusters 3, 4, 5 and 9 across samples (Additional File 1: Table S9). However, only cluster 4 remained significant after correcting for baseline case enrichment and multiple testing, and only when combining PTV, misB, and misA variants (Fisher’s exact test, OR=14.8, p=8.1x10^-4^), (Additional File 1: Table S10). Genes in cluster 4 were strongly associated with the nucleosome (GO:0000786, GO p= 4.73x10^-40^), with 19 of the 23 queried genes contributing to the signal including *H3F3A*, HIST1H4H, *KAT6A* and *KAT6B*, which each carried *de novo* variants in cases. The nucleosome is a complex in which DNA is wrapped around histone proteins, forming the fundamental unit of chromatin. Mutations in nucleosome-associated genes have been implicated in dozens of developmental disorders and are associated with a broad spectrum of craniofacial anomalies, including OFC [69,70]. Overall, this network analysis highlights candidate genes for future study that are biologically connected with potential relevance to OFC etiology.

### Evidence Based Classification of Novel OFC Candidate Genes

We examined the interrelatedness among our newly identified OFC candidate genes from the above analyses (gene set enrichment, TADA, protein network) and after excluding the 593 genes with prior established associations to OFC, we identified 231 genes with at least one deleterious variant (PTV or misB) in our cohort (Fig. 4A). The most frequent combination of gene categories involved constrained genes (n = 84) absent of other categories and constrained OMIM autosomal dominant genes (n = 58). Of the 231 candidate genes, 32 harbored at least two deleterious variants (Fig. 4B), which we systematically reviewed in the literature to further evaluate their association with OFC and classify them based on the strength of existing evidence. Accordingly, we grouped the genes into three categories: (i) **Supported in literature**, in which an association with OFC has already been reported; among these we identified 16 genes with prior support in the literature (e.g. *ZFHX3*, *GRHL2*). (ii) **Possible phenotype expansion of known syndrome**, where the gene is associated with a known syndrome but has not previously been linked to OFC or PA in our review: *CACNA1E*, *CDK13*, *CSNK2B*, *EIF3A, KIF1A*, *NR2F2*, *SOS1,* and *SUV420H1*. (iii) **Newly emerging genes**, defined as genes for which we found no supporting evidence in the literature; these include *ARIH1, BRSK2, ELAVL3, KPNA1, NLGN2, SF3B3, SLC4A4,* and *TM9SF3* (Fig. 4B, Additional File 1: Table S11).

**Fig. 4.**
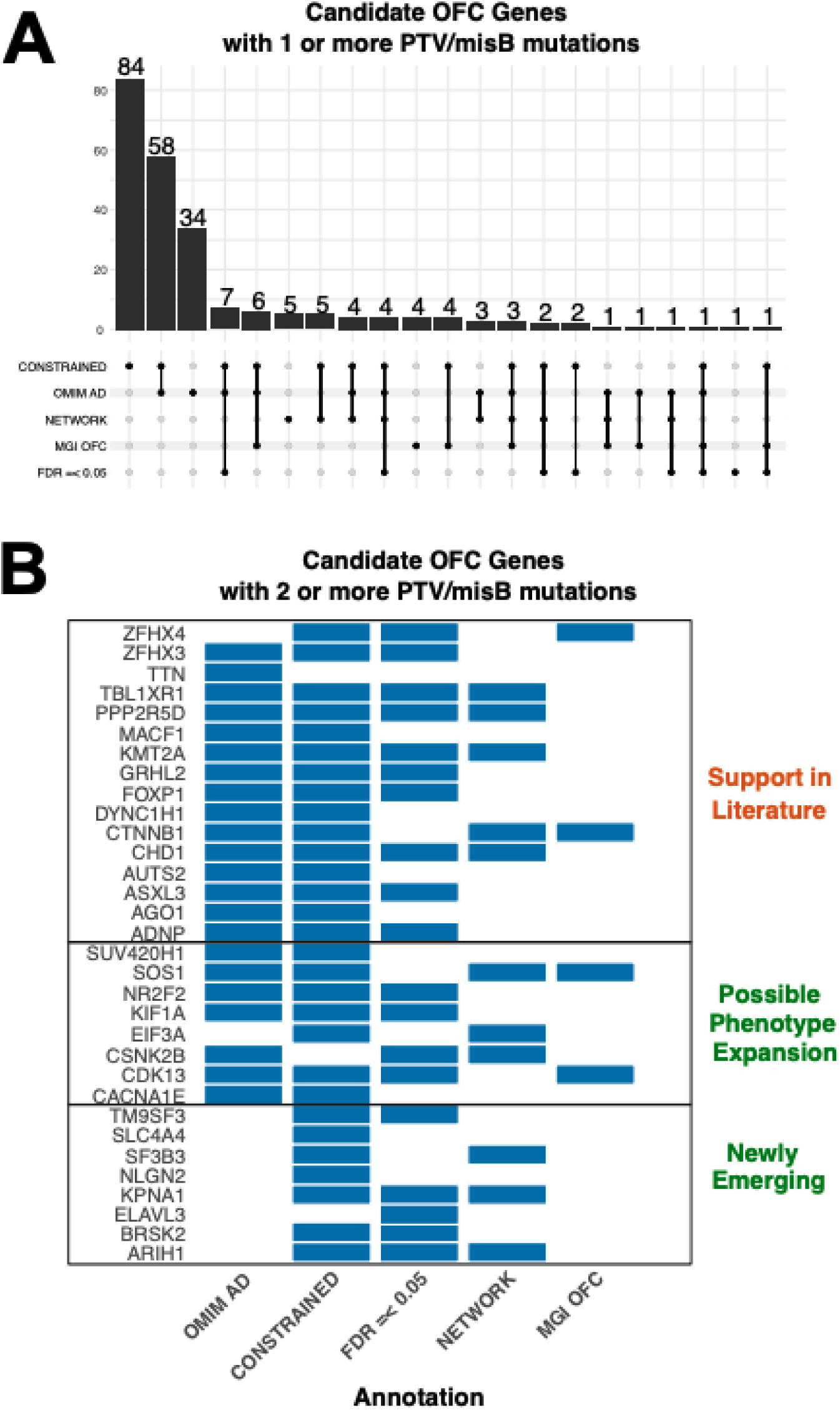
Overlap of OFC Candidate Genes Across Analyses. **(A)** Upset plot summarizing the number of candidate OFC genes (excluding n = 593 established OFC genes) identified under different annotation categories. Categories include genes supported by mouse OFC (MGI OFC), genes within OFC protein interaction networks, genes associated with OMIM AD, genes under mutational constraint and significant genes (FDR ≤ 0.05) from our gene association analysis. Counts are shown for genes with ≥1 PTV or misB mutations across cases. **(B)** Annotation categories shown across candidate genes found harboring ≥2 PTV or misB case variants. Genes are grouped based on prior various levels of support in the scientific literature.

## Discussion

This work represents a large systematic assessment of coding *de novo* variants in 2,497 trios with an affected proband with OFC or PA. We evaluated the global enrichment of OFC relative to control trios and, similar to past studies [4,13], observed significant enrichment of both PTVs and highly deleterious missense variants across syndromic and nonsyndromic OFCs, with the exception of nonsyndromic cleft lip, which has been shown to be less influenced by strong effect *de novo* mutations [71,72]. We further parsed biologically relevant gene sets and showed further enrichment across known OFC genes, evolutionarily constrained genes, Mendelian disorder genes absent an established OFC association, and mouse candidate genes. To complement these findings, we applied an unbiased gene discovery approach using the TADA method, which identified 39 significant genes at an FDR of 0.05 and 16 at a more rigorous FDR<0.001. We then constructed a PPI network seeded with established OFC genes, revealing a significant enrichment of *de novo* variants in cases among high-order interactors with at least 5 connections to a seed gene. Based on these results, we built a high-confidence network of 533 genes, which clustering analysis identified associations with 81 distinct biological functions, including 10 categories containing at least 10 genes, spanning diverse pathways such as chromatin organization and transcriptional regulation. Across all analyses, we identified 231 candidate genes; however, because OFC and PA are relatively common phenotypes, it is often difficult to determine from individual cases whether these features are truly part of the associated phenotypic spectrum or coincidental findings, whereas restricting analyses to genes with at least two deleterious *de novo* variants in our cohort identified 32 higher-confidence candidate genes. A number of these had orthogonal support in the literature (Additional File 1: Table S11), and we therefore avoid making any claims of novelty, instead we aim to further strengthen their candidacy, facilitating their incorporation into diagnostic testing and improving prioritization for functional studies.

We took a maximal approach in this study combining both syndromic and nonsyndromic cases, as well as incorporating PA into our data. We hypothesized the architecture of these disorders are similar enough that by combining them, we would gain power for gene discovery, which is an approach previously successful in other congenital anomaly and neurodevelopmental disorders genomic studies [22,73–75]. We found this approach to be highly successful in this case, identifying nine additional genes at an FDR < 0.05 compared to restricting the study to separate entities (syndromic vs nonsyndromic). Moreover, the inclusion of 417 cases with PA in the absence of OFC, a phenotype not traditionally incorporated into OFC studies, enhanced gene discovery, yielding eight additional candidate genes. Among our strongest candidate genes harboring two or more deleterious mutations in the cohort (n = 32), we found 34.4% to include at least one PA case and one OFC case, underscoring a shared genetic architecture between these phenotypes and suggesting that, in some instances, PA may represent a milder manifestation along the cleft palate spectrum. Expanding on this work, a future step could incorporate other related congenital anomalies, such has recently been performed between OFC and congenital heart defects [76].

A limitation of this study, as with research on any isolated anomalies, is the potential for ascertainment bias, since cases are often collected at a young age before certain comorbidities (e.g., neurodevelopmental) have manifested. We investigated the potential impact of this among our nonsyndromic OFCs with mutations in established OFC syndromic genes (n=20, OMIM AD genes) and observed 80% are associated with at least one major neurodevelopmental comorbidity, such as epilepsy, autism, intellectual disability or developmental delay indicating that some cases classified as nonsyndromic may later develop additional clinical features. Specifically for genes like like *CSNK2B*, where genetic testing of an individual with an OFC could identify a pathogenic variant prior to onset of seizures and would be valuable for medical management and surveillance. These findings therefore underscore the importance of longitudinal clinical follow-up for patients to accurately distinguish nonsyndromic from syndromic OFC.

## Conclusions

In summary, we performed a robust analysis of a cohort of 2,497 trios with probands affected by OFC or PA, encompassing both syndromic and nonsyndromic cases. We identified a significant enrichment of *de novo* mutations in these cases when compared against 5,515 control trios. These *de novo* mutations were enriched in a variety of biologically relevant gene sets (e.g. genes under evolutionary constraint), reinforcing their importance in the genetic architecture of OFC. Gene association analysis identified 39 genes at an FDR<0.05 and a network analysis found 309 high-order interactors that largely clustered into ten distinct biological pathways, with genes associated with the nucleosome showing a strong enrichment among cases in our cohort. Finally, we aggregated results across all analyses to identify 231 candidate genes, 32 of which harbored at least two deleterious variants in our cohort, including 16 with prior support in the literature, eight representing potential phenotypic expansions of known syndromes, and eight with little to no existing support. Overall, this study provides a comprehensive view of the genetic etiology underlying OFC by integrating multiple analytic approaches, highlighting both established and emerging candidate genes, and underscoring the value of large-scale, harmonized datasets for gene discovery and improved biological insight.

## Data Availability

All sequencing data is publicly available with sources highlighted in Additional file: Table S1.

## List of abbreviations

AC: Allelic Count
AD: Allelic Depth
AD: Autosomal Dominant
AF: Allelic Frequency
ASD: Autism Spectrum Disorder
BF: Bayes Factor
CL: Cleft Lip
CLP: Cleft Lip with Cleft Palate
CNV: Copy Number Variations
CP: Cleft Palate
CPX: Complex SV
CTX: Reciprocal Translocations
DEL: Deletions
DUP: Duplications
ES: Exome Sequencing
FDR: False Discovery Rate
GATK: Genome Analysis Toolkit
GD: Genomic Disorder
gnomAD: Genome Aggregation Database
GQ: Genotype Quality
GS: Genome Sequencing
GWAS: Genome wide association studies
HPO: Human Phenotype Ontology
IBD: Identity-by-descent Indels Insertions/deletions
INS: Insertions
INV: Invertions
IQR: Interquartile Range
LOEUF: Loss-of-function Observed/Expected Upper bound Fraction
LoF: Loss-of function
MCL: Markov Cluster Algorithm
MGI: Mouse Genome Informatics
PMC: Missense Deleteriousness Metric
OFC: Orofacial Cleft
OMIM: Online Mendelian Inheritance in Man
PA: Palate Abnormality
PCA: Principal Component Analysis
POBINDS: Poirier-Bienvenu Neurodevelopmental Syndrome
PPI: Protein–Protein Interaction
PTV: Protein-Truncating Variant
PURF: Positive-Unlabeled Random Forest
Q95: 95th percentile
QC: Quality Check
SNVs: Single Nucleotide Variants
SV: Structural Variant
VCF: Variant Call Format
VEP: Variant Effect Predictor
VQSLOD: Variant Quality Score Recalibration

## Declarations

### Ethics approval and consent to participate

All analyses were conducted in accordance with the Mass General Brigham (MGB) IRB protocol #2019P003282.

### Consent for publication

Not Applicable.

### Availability of data and materials

All sequencing data is publicly available with sources highlighted in Additional file: Table S1.

### Authors’ contributions

Study design and concept: N.E.K., A.S.-J., M.T., M.L.M., E.J.L.-C., H.B., Sample collection and data sharing: S.W.C., K.R.R., A.A.A., K.K.D.P., L.J.J.G., M.A.E., W.L.A., C.J.B., C.D.P., F.A.P., G.L.W., J.T.H., L.M.M.U., N.M., J.R.S., S.M.W., J.C.M., T.H.B., A.B., M.L.M., E.J.L.-C., Variant Calling and Computational analysis: N.E.K., A.S.-J., E.S., S.C., K.R., A.S.L., X.Z., J.F., A.C.T., E.J.L.-C., H.B. Project oversight: H.B., M.E.T. Primary Manuscript writing and figure generation: N.E.K., A.S.-J., H.B.

## Acknowledgments

We gratefully acknowledge the participants, their families, and our colleagues, whose contributions and support were essential to this work, including those from the following studies: (1) The DDD study presents independent research commissioned by the Health Innovation Challenge Fund [grant number HICF-1009-003], a parallel funding partnership between Wellcome and the Department of Health, and the Wellcome Sanger Institute [grant number WT098051]. The views expressed in this publication are those of the author(s) and not necessarily those of Wellcome or the Department of Health. The study has UK Research Ethics Committee approval (10/H0305/83, granted by the Cambridge South REC, and GEN/284/12 granted by the Republic of Ireland REC). The research team acknowledges the support of the National Institute for Health Research, through the Comprehensive Clinical Research Network. This study makes use of DECIPHER (http://www.deciphergenomics.org), which is funded by Wellcome [grant number WT223718/Z/21/Z]. See Nature PMID: 25533962 or www.ddduk.org/access.html for full acknowledgement. (2) The Undiagnosed Diseases Network (UDN) is supported by the National Institute of Neurological Disorders and Stroke of the National Institutes of Health under award numbers U01NS134348, U01NS134357, U01NS134352, U01NS134350, U01NS134358, U01NS134356, U01NS134353, U01NS134355, U01NS134351, U01NS134349, U01NS134354 and U2CNS132415. Earlier phases of the UDN were supported in part by the Intramural Research Program of the National Human Genome Research Institute and by the National Institutes of Health Common Fund, through the Office of Strategic Coordination/Office of the NIH Director under award numbers U01HG007674, U01HG007703, U01HG007709, U01HG007672, U01HG007690, U01HG007708, U01HG007530, U01HG007942, U01HG007943, U01TR002471, U54NS093793, U54NS108251, U01HG010215, U01HG010233, U01HG010217, U01HG010230, U01HG010219, and U01TR001395. The content is solely the responsibility of the authors and does not necessarily represent the official views of UDN investigators or the National Institutes of Health. The datasets used for the analyses described in this manuscript were obtained from dbGaP at http://www.ncbi.nlm.nih.gov/gap through dbGaP accession number phs001232. (3) This work is supported by generous donors to the Children’s Mercy Research Institute and the Genomic Answers for Kids Program (GA4K) at Children’s Mercy, Kansas City accessible at phs002206. (4) National Institutes of Health (NIH) support was provided in part by a grant from the National Human Genome Research Institute and the National Heart, Lung, and Blood Institute (1U54HG006542) to Dr. David Valle for the Baylor Hopkins Center for Mendelian Genomics program. The dataset(s) used for the analyses described in this manuscript was obtained from the database of Genotype and Phenotype (dbGaP) found at http://www.ncbi.nlm.nih.gov/gap through dbGaP accession number phs000711. (5) These analyses included data generated as part of the GREGoR Consortium, which is supported by the National Human Genome Research Institute through: U01HG011762, U01HG011744, U01HG011755, U01HG011758, U01HG011745, and U24HG011746. (5) The results analyzed and published here are based in part upon data generated by Gabriella Miller Kids First (GMKF) Pediatric Research Program projects phs001420, phs001997, phs001168, phs002595 and which can be accessed from the Kids First Data Resource Portal (https://kidsfirstdrc.org/) with dbGaP approval.

## Supplementary Information

Additional File 1.xlsx

Additional File 1 Supplementary Tables: Table of contents, **Table S1**: Cohort definition. **Table S2**: Cohort Phenotype breakdown. **Table S3**: Full gene list with annotation. **Table S4**: Enrichment table (case vs control). **Table S5**: Enrichment Table (case target vs case background gene). **Table S6**: TADA genes. **Table S7**: Network genes and clusters. **Table S8**: Pathways. **Table S9**: Network enrichment table (case vs control). **Table S10**: Network enrichment table (case target vs case background gene). **Table S11**: OFC candidate genes.

Additional File 2.doc

Additional File 2 Supplementary Figures: **Fig. S**1: Classification of OFC Patients. **Fig. S2**: Bar plots illustrating the distribution of syndromic and non-syndromic OFC groups. **Fig. S3**: De novo mutation rates by variant type. **Fig. S4**: Assessment of enrichment of first-order interactor genes by connectivity.

Additional File 3.doc

Additional File 3 Supplementary Methods.

