## Additional file 2 for "Comprehensive analysis of *de novo* variants across 2,497 orofacial cleft trios reveals novel genetic drivers of disease"

### **Supplemental Figures**

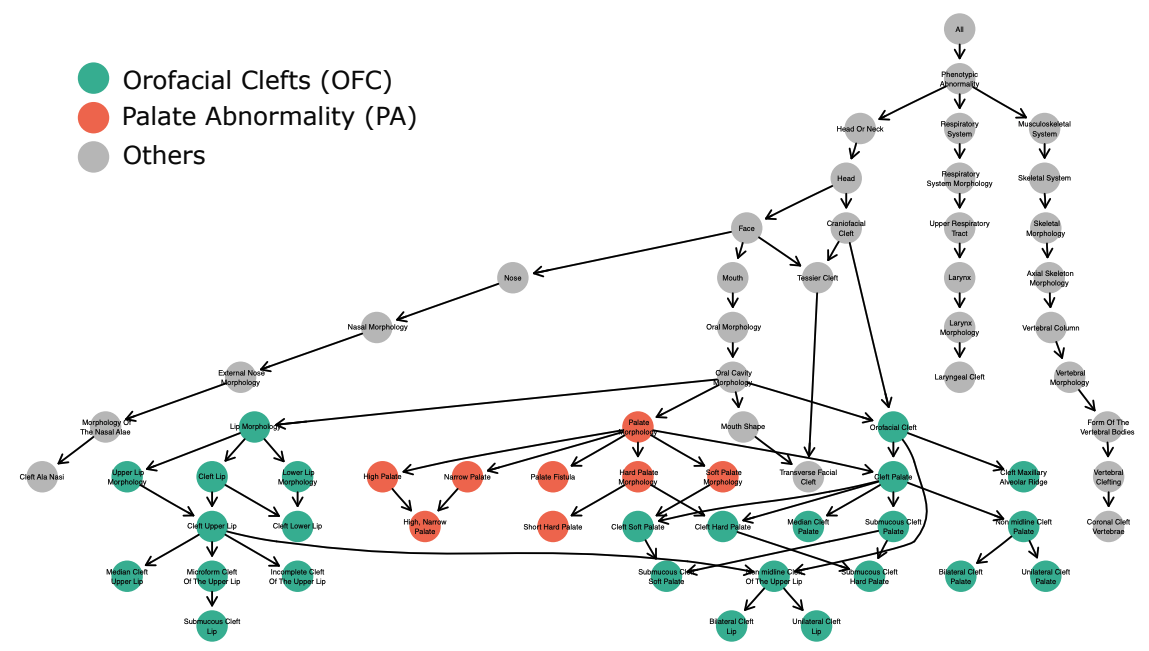

**Fig. S1: Classification of Orofacial Cleft (OFC) Patients:** Sample phenotypes were collected as Human Phenotype Ontology (HPO) terms [1]. Each OFC case was classified by subtype and further categorized as isolated or syndromic based on the presence of additional anomalies. We extracted HPO IDs using the hpo.obo file via the ontologyIndex library in R software [2]. HPOs related to orofacial clefting (Green) and palate anomalies (Red) were extracted. OFC subtypes were assigned using a hierarchical classification scheme with samples based on cleft palate and cleft lip HPO assignment. Samples without an orofacial cleft but still having a palate abnormality (PA) were classified as PA and samples with only generic OFC terms were categorized as unspecified OFC.

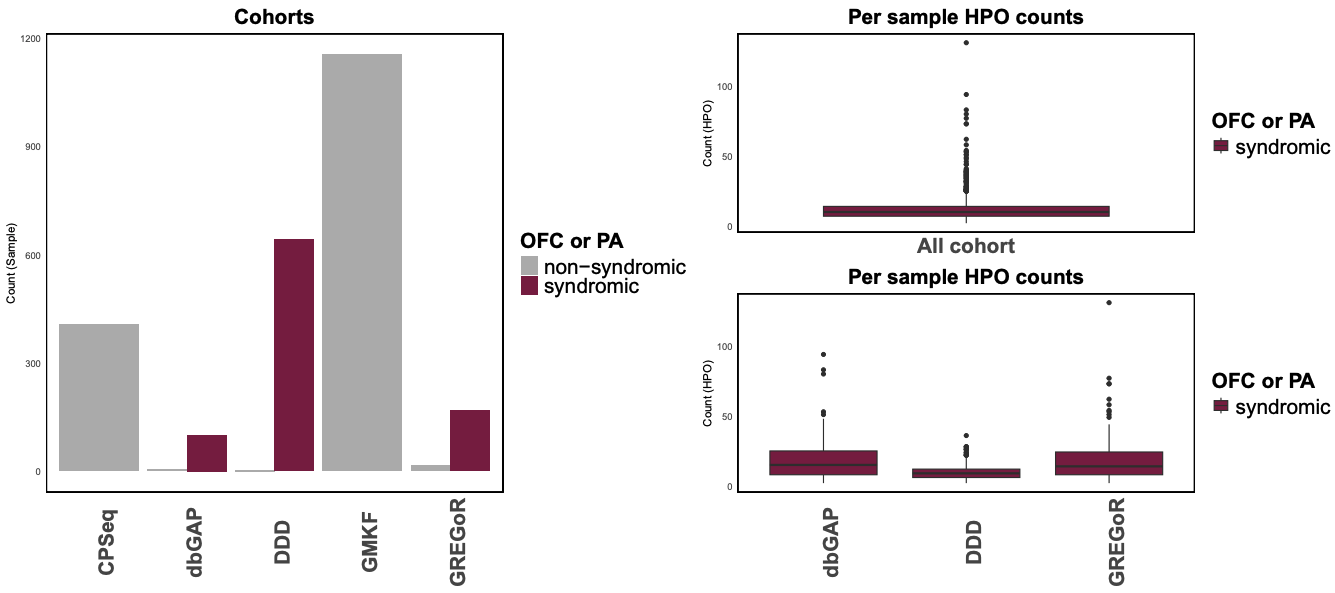
**Fig. S2: Bar plots illustrating the distribution of syndromic and non-syndromic OFC groups.** Right panel: the y-axis shows the number of samples per source cohort, stratified by syndromic status (syndromic in purple and non-syndromic in gray). Left panel: the y-axis shows the number of HPO terms per sample for syndromic cases, shown for the entire cohort (top) and further stratified by source cohort (bottom).

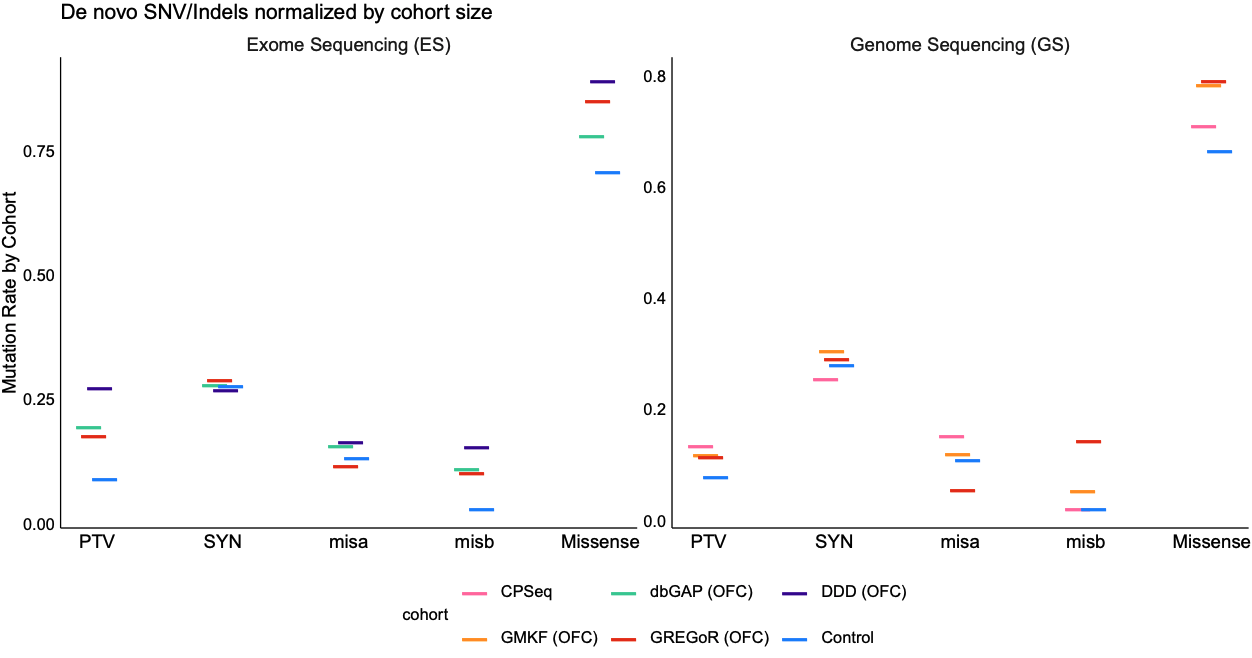

**Fig. S3: *De novo* mutation rates by variant type**. The y-axis shows the de novo mutation rate per cohort, with ES data on the left and GS data on the right. The variants are broken in by functional categories; protein truncating variants (PTV), synonymous variants (SYN) and missense variants; which further classified into functional categories based missense deleteriousness metric (MPC) metric [3]; as misB (highly disruptive, MPC ≥ 2) and misA (moderately disruptive, 1 ≤ MPC < 2).

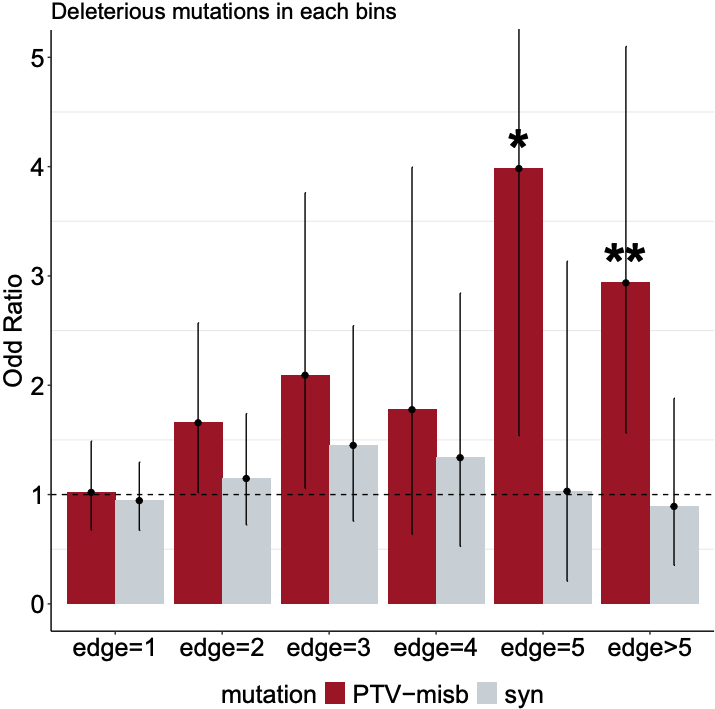

**Fig. S4: Assessment of enrichment of first-order interactor genes by connectivity.** Bar plots show odds ratios for mutation enrichment across network connectivity bins (edge = 1, 2, 3, 4, 5, and >5), where an edge represents a connection to an established OFC gene. Deleterious mutations (PTV + misB; red) and synonymous mutations (syn; gray) are shown separately. The dashed line marks an odds ratio of 1 (no enrichment). Asterisks highlight statistically significant enrichment after Bonferroni correction.
