## Additional file 3 for "Comprehensive analysis of *de novo* variants across 2,497 orofacial cleft trios reveals novel genetic drivers of disease"

### **Supplementary Methods**

**De Novo Variant Filtering for Exome Sequencing (ES)**

Variants were first filtered based on VCF filter status requiring a “PASS” and excluding calls with extremely low or high sequencing depth (DP < 7 or DP > 1000). Next, homozygous reference (0/0) calls were excluded if genotype quality (GQ) was below 25. Homozygous variant (1/1) calls were removed if the phred-scaled likelihoods (PL) for reference genotype (0/0) was below 25, and heterozygous (0/1) calls were removed if PL(0/0) was below 25. This is followed by sex-specific genotype filtering accounting for hemizygous and pseudoautosomal regions (PAR). In females, any low depth calls (<10) or any calls on the Y chromosome were removed. In males, any low depth calls (<10) at autosomal and PAR were removed and heterozygous calls in hemizygous regions were also removed. For heterozygous genotypes, Hail’s binom_test module (https://github.com/hail-is/hail/) was used to compute a two-sided binomial allele balance p-value (pAB), using the allelic depth (AD) of the alternate allele to represent the probability of the expected AB value of 0.5. The following filters were then applied sequentially to keep only informative heterozygous calls. Heterozygous calls with allele balance (AB) below 0.25 and alleles with pAB < 1×10⁻⁹ were excluded. Heterozygous calls with a sum of AD less than 90% of the depth were removed. Similarly, homozygous calls with alternate AD less than 90% of the depth were removed. We then calculated the call rate, taking into account male hemizygous calls, such that non-PAR chrY variants had a call rate of (males with a defined genotype) / (number of males), and non-PAR chrX variants had a call rate of (males with a defined genotype + 2 * females with a defined genotype) / (number of male samples + 2 * number of female samples). Otherwise, the call rate for a given variant was defined as (number of samples with a genotype) / (number of samples). Variants with a call rate <0.8 were excluded. We computed the p-value of the Hardy-Weinberg equilibrium test (pHWE) and removed variants with a pHWE <1×10⁻¹². After these quality filters, we removed low quality calls with GQ <25. Population frequency priors collected by non-neuro gnomAD allele frequencies (exomes v2.1.1).

**De novo Variant Filtering for Genome Sequencing (GS)**

SNV/Indels with cohort Allelic Count ≥ 20 and loci in low-complexity regions (LCRs) were removed. The loci where parents are homozygous reference and probands are heterozygous were retained. For SNVs, variants with PASS for Variant Quality Score Recalibration (VQSR) were retained. Thresholds for quality metrics differed for SNVs and indels. For SNVs, QUAL ≥150, SOR ≤2.5, ReadPosRankSum ≥ −1.4, QD ≥3.0, and MQ ≥50; while for indels, QUAL ≥150, SOR ≤3, ReadPosRankSum ≥ −1.7, QD ≥4.0, and MQ ≥50. Additionally, calls were retained if they met any of the following criteria: for SNVs, (1) parental GQ ≥30 and AB ≤0.05 or (2) proband GQ ≥99 and 0.22 ≤ AB ≤ 0.78; for indels, (1) DP ≥16, GQ ≥30, and AB ≤0.05 or (2) proband GQ ≥99 and 0.2 ≤ AB ≤ 0.8. Variants were further filtered for a mean GQ ≥50. Following these quality filters, outlier samples were dropped by defining a maximum threshold for the number of variants per proband (restricted to those passing sex and relatedness QC) and removing any sample exceeding this threshold.

We trained a Positive-Unlabeled Random Forest (PURF) model to filter de novo. Two separate models were implemented, one for variant-level features and one for sample-level features. The initial positive labels were defined as ultra-rare variants present in one parent but absent in the proband for the variant-level model, and as ultra-rare inherited variants for the sample-level features, to factor in AC-based bias in quality metrics. The variant-level model used MQ, FS, BaseQRankSum, SOR, LEN, ReadPosRankSum, DP, QD, and VQSLOD as features. The sample-level model used GQ_parent, AB_sample, DPC_sample, DPC_parent, PL_sample_0.0, and PL_sample_1.1. In addition, the PURF model was implemented on indels with length ≤3 and length >3 separately. The ultra-rare variant call set was curated using the same quality criteria applied to de novo variants and was further filtered to drop variants with missing VQSLOD or VQSLOD values below −10. For cohorts containing a high number of ultra-rare variants, we downsampled the call set to approximately align with the count of putative de novo variants. The PURF model was implemented using the PU bagging method with n_bags=10 and n_estimators=1000, and we evaluated the model via k-fold cross validation with k=5.

**Annotation of variants**

Final SNV/Indel callset were annotated for predicted functional consequence using the Ensembl Variant Effect Predictor (VEP) v112 [1], with the AlphaMissense [2] and EVE plugins [3]. Additional variant annotations were generated using a custom pipeline implemented in Hail, integrating data from ClinVar VCF version 03/06/2025 [4], CADD v1.6 [5], EIGEN [6], LOEUF scores [7] from gnomAD v2.1.1, MPC scores v1 [8], allele frequencies from gnomAD exomes and genomes, REVEL [9], and precomputed SpliceAI scores [10]. Functional consequences were prioritized per variant and gene, and the most severe consequence was selected using an adaptation of the gnomAD Python API module gnomad.utils.vep.process_consequences.

**Generating MGI OFC Gene list**

We also collected genes associated with OFC phenotypes in mice using the Mouse Genome Informatics (MGI) database[11], where Mouse Phenotype (MP) definition includes “cleft” or “palate” and then removing any terms not specifically related to an OFC. These genes were then mapped to their human homologs, which resulted in a total of 362 genes, which reduced to 349 genes when focusing on our list of autosomal coding genes.
